# Detailed patient-individual reporting of lymph node involvement in oropharyngeal squamous cell carcinoma with an online interface

**DOI:** 10.1101/2021.12.01.21267001

**Authors:** Roman Ludwig, Jean-Marc Hoffmann, Bertrand Pouymayou, Grégoire Morand, Martina Broglie Däppen, Matthias Guckenberger, Vincent Grégoire, Panagiotis Balermpas, Jan Unkelbach

## Abstract

**Purpose/Objective:** Whereas the prevalence of lymph node level (LNL) involvement in head & neck squamous cell carcinomas (HNSCC) has been reported, the details of lymphatic progression patterns are insufficiently quantified. In this study, we investigate how the risk of metastases in each LNL depends on the involvement of upstream LNLs, T-category, HPV status and other risk factors.

**Materials/Methods:** We retrospectively analyzed patients with newly diagnosed oropharyngeal HNSCC treated at a single institution, resulting in a dataset of 287 patients. For all patients, involvement of LNLs I-VII was recorded individually based on available diagnostic modalities (PET, MR, CT, FNA) together with clinicpathological factors. To analyze the dataset, a web-based graphical user interface (GUI) was developed, which allows querying the number of patients with a certain combination of co-involved LNLs and tumor characteristics.

**Results:** The full dataset and GUI is part of the publication. Selected findings are: Ipsilateral level IV was involved in 27% of patients with level II and III involvement, but only in 2% of patients with level II but not III involvement. Prevalence of involvement of ipsilateral levels II, III, IV, V was 79%, 34%, 7%, 3% for early T-category patients (T1/T2) and 85%, 50%, 17%, 9% for late T-category (T3/T4), quantifying increasing involvement with T-category. Contralateral levels II, III, IV were involved in 41%, 19%, 4% and 12%, 3%, 2% for tumors for tumors with and without midline extension, respectively. T-stage dependence of LNL involvement was more pronounced in HPV negative than positive tumors, but overall involvement was similar. Ipsilateral level VII was involved in 14% and 6% of patients with primary tumors in the tonsil and the base of tongue, respectively.

**Conclusions:** Detailed quantification of LNL involvement in HNSCC depending on involvement of upstream LNLs and clinicopathological factors may allow for further personalization of CTV-N definition in the future.

## Introduction

Head & neck squamous cell carcinomas (HNSCC) spread though the lymphatic system of the neck and form metastases in regional lymph nodes. Therefore, the target volume in radiotherapy of HNSCC patients includes, in addition to the primary tumor, parts of the lymph drainage volume [1], [2]. The nodal gross tumor volume (GTV-N) contains detectable macroscopic lymph node metastases, while the elective clinical target volume (CTV-N) contains parts of lymph drainage volume that is at risk of harboring microscopic tumor, i.e., occult metastases that are not yet visible with current imaging techniques.

GTV-N definition is primarily performed through imaging techniques (PET-CT/MRI, MRI, or CT) as well as fine needle punctures (FNA). Imaging criteria for lymph node metastases include size, round rather than oval shape, central necrosis, and FDG uptake as summarized by Biau et al [1]. Goel et al. gives an overview over clinical practice in PET/CT for the management of HNSCC [3]. However, all imaging techniques have finite sensitivity and specificity [4]–[6], i.e. they fail to detect small metastases or may incorrectly identify suspicious lymph nodes as tumor.

For standardized reporting of the location of lymph node metastases as well as delineation of the CTV-N, the lymph drainage system of the neck is divided into anatomically defined regions called lymph node levels (LNL) [7]. CTV-N definition amounts to the decision which LNLs to include into the elective CTV-N and is based on international consensus guidelines. Such guidelines were first published in 2000 by Grégoire et al and have been updated in 2006, 2014 and 2019 [1], [7]–[9]. Current recommendations for the selection of lymph node levels in oropharyngeal cancer can be found in Table 2 of the 2019 published delineation guideline by Biau et al [1]. Current guidelines are primarily based on the prevalence of LNL involvement for a given primary tumor location, i.e., the percentage of patients diagnosed with metastases in each level. It is recommended that the elective CTV-N includes all LNLs that are involved in 10–15% of patients or more. Patients are primarily stratified by primary tumor location. For example, tumors of the soft palate, the posterior pharyngeal wall and the base of tongue show lymph node metastases on both sides via crossing lymph vessels. For this reason, even for lateralized tumors of these localizations, bilateral neck treatment is recommended. However, the lymphatic drainage of the tonsil is mainly unilateral, therefore an ipsilateral irradiation is recommended for lateralized low T-category (T1/T2) tumors (at least up to lymph node stage N2a). Volume-reduced elective nodal irradiation has been or is being investigated in several trials [10], [11]

While the general patterns of lymph drainage in the neck is understood and prevalence of LNL involvement has been reported in the literature [8], [12], [13], [14], the details of progression patterns in oropharyngeal HNSCC are poorly quantified. How much does the risk of level IV involvement increase depending on whether levels II and III harbors macroscopic metastases? How much does the risk of involvement increase for late versus early T-category? Are progression patterns different for HPV positive versus HPV negative tumors? Answering these questions quantitatively may allow for further personalizing CTV-N definition based on an individual patient’s clinical presentation at the time of diagnosis.

The basis for better quantification of LNL involvement are detailed datasets of HNSCC patients for whom involvement is reported for each individual LNL together with tumor and patient characteristics. For example, to answer the question, how much the risk in level IV increases depending on the involvement of upstream levels II and III, it is insufficient to only report prevalence of LNL involvement in levels II, III, and IV. Instead, one needs to know in how many patients a certain combination of involvement is observed, i.e., how often are levels II, III, IV all involved simultaneously, versus how often is only level II and III involved but not IV, and so on. The contributions of this work are:

- We provide a dataset of lymphatic progression patterns in 287 oropharyngeal HNSCC patients treated at our institution in whom involvement of LNLs together with tumor characteristics are reported on a patient-individual basis.
- To visualize and explore the complex dataset, a graphical user interface is provided that allows the user to query the number of patients who were diagnosed with a specific combination of simultaneously involved LNLs and tumor characteristics.

We hope that this work provides the basis for collecting large multicenter datasets of lymphatic progression patterns, which can then inform future guidelines on further personalized CTV-N definition.

## Material & Methods

### Data curation

We included patients diagnosed with oropharyngeal squamous cell carcinoma (primary diagnosis) between 2013 and 2019 and treated at the department of radiation oncology and/or head and neck surgery of the University Hospital Zurich (USZ). Patients with prior radiotherapy or surgery to the neck were excluded, resulting in a dataset of 287 patients. Specific subsites of oropharyngeal cancer included the base of tongue, the tonsils as well as the oropharyngeal side of the vallecula and the posterior or lateral wall of the oropharynx. Patient information consisted of the date of birth, gender, the date of the 1^st^ histological confirmation of the tumor, the performed treatment (surgery with neck dissection prior to RT/RCHT *vs*. surgery only *vs*. definitive radio(chemo)therapy), risk factors such as nicotine abuse and HPV-status (p16 pos/neg), the TNM-classification (UICC 7^th^ edition until 2017, 8^th^ edition since 2017), the position of the primary tumor (left/right neck) as well as positive *vs*. negative midsagittal plane extension. Further details are described in the Appendix A: Details on data curation.

The analysis of the lymphatic spread included levels Ia, Ib, IIa, IIb, III, IV, V, VII and was performed separately for the diagnostic imaging modalities available for a patient (FDG PET-CT, FDG PET-MRI, MRI, CT) as well as FNA and radiotherapy planning CT if available. This was performed by 2 experienced radiation oncologists by reviewing radiology and pathology reports together with the diagnostic images. Criteria for considering a lymph node as malignant followed the description in Biau et al [1] and are described in detail in appendix A below.

### Data base

The full dataset is available as a CSV-file on GitHub: https://github.com/rmnldwg/lydata/tree/main/2021-oropharynx.

### Graphical user interface

We developed an online GUI based on the Django framework [15] and provide it to explore the dataset. It allows the user to conveniently determine the number of patients that show a particular combination of co-involved LNLs and tumor characteristics. The GUI is available at https://2021-oropharynx.lyprox.org, its source code under MIT license is available on GitHub: https://github.com/rmnldwg/lyprox. Documentation is provided within the GUI along with videos demonstrating its use.

## Results

In this section, we summarize selected findings. To that end, LNL involvement based on CT, MRI, PET and FNA was converted into a consensus decision via a logical OR, i.e., a LNL is considered involved if it was positive for one of these 4 modalities. However, only a small fraction of the information contained in the full dataset can be summarized here in tables. Thus, the interested reader is encouraged to explore the full dataset directly.

### Graphical user interface

Figure 1 shows the GUI for analyzing the dataset. The main patient characteristics (smoking status, HPV status, and primary treatment) are shown on the top left. In the bottom-left panel, the user can specify characteristics of the primary tumor. In the example, the user considers all subsites combined, but restricts the patient selection to late T-category tumors (T3/T4) that extend over the midsagittal plane. On the top-right, the user selected the diagnostic modalities CT, MR, PET and FNA, which are connected via a logical OR, i.e., a lymph node level is considered as involved for a patient if it was considered positive for at least one of the modalities available for that patient. The main panel shows the involvement of LNLs. In the example, the user restricts the selection to patients with positive ipsilateral level III while all other levels are unspecified. In total, 38 out of 287 patients meet these selection criteria. The main panel now displays the number of patients with involvement of the other levels. E.g., 21 patients have contralateral level II involved, and 12 patients ipsilateral level IV.

**Figure 1:**
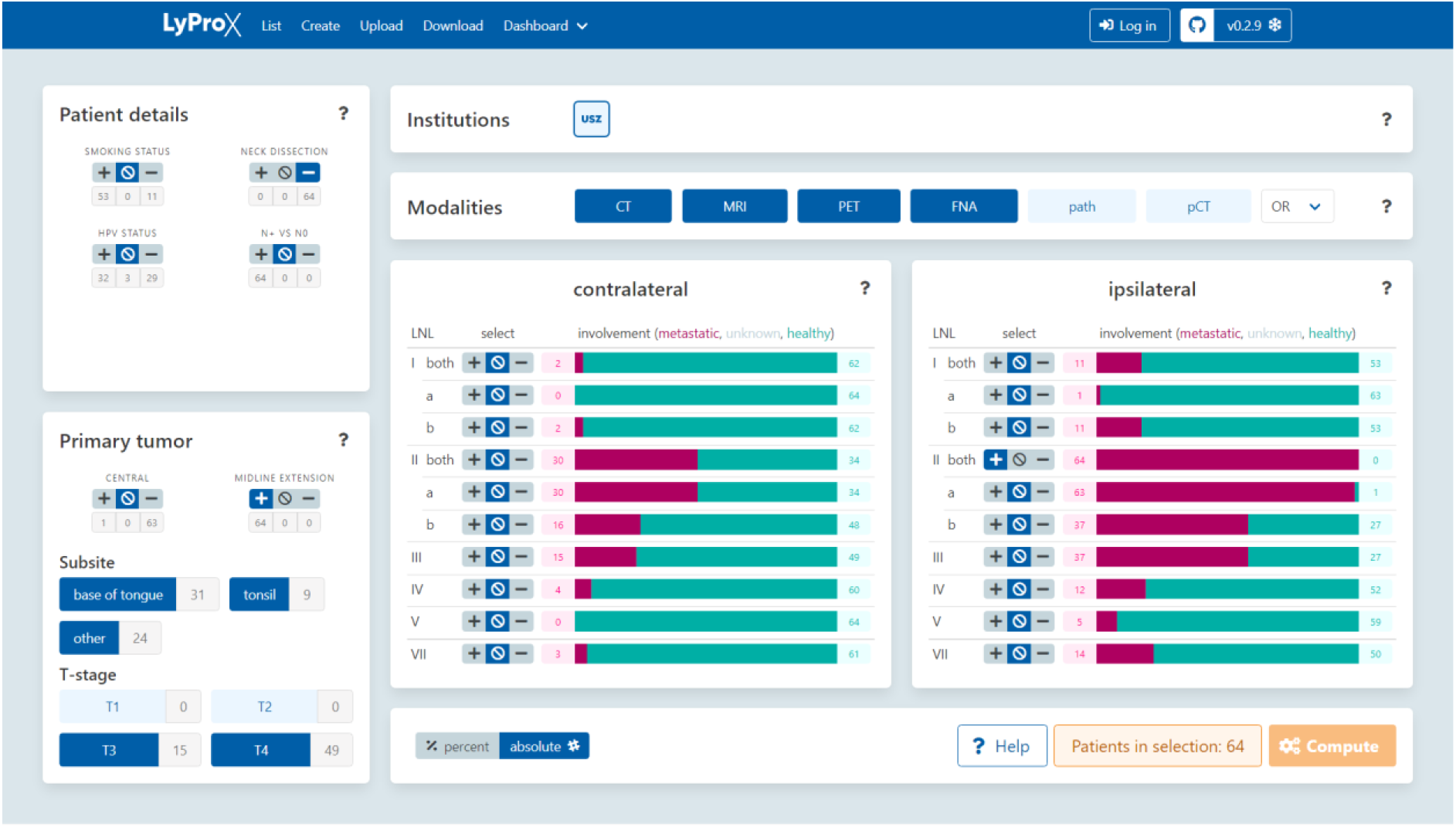
GUI to analyze lymphatic metastatic progression patterns.

### Prevalence

Overall prevalence of lymph node level involvement is reported in Table 1 and visualized in Figure 2a.

**Table 1:** Prevalence of LNL involvement for the whole patient cohort (all) and stratified according to early (T1/T2) versus late (T3/T4) T-category and HPV positive (HPV+) versus HPV negative (HPV-) tumors. For each LNL, the first column indicates the number of patients showing involvement in the level, the second column the percentage of positive patients in the respective group.

|  | n | I | II | III | IV | V | VII |
| --- | --- | --- | --- | --- | --- | --- | --- |
| **ipsi all** | 287 | 30 10% | 232 81% | 113 39% | 31 11% | 16 6% | 30 10% |
| **T1/T2** | 150 | 10 7% | 118 79% | 48 32% | 10 7% | 4 3% | 12 8% |
| **T3/T4** | 137 | 20 15% | 114 83% | 65 47% | 21 15% | 12 9% | 18 13% |
| **HPV+** | 181 | 20 11% | 155 86% | 73 40% | 20 11% | 12 7% | 18 10% |
| **HPV-** | 96 | 8 8% | 69 72% | 37 39% | 11 11% | 4 4% | 12 13% |
| **HPV+, T1/T2** | 100 | 7 7% | 86 86% | 35 35% | 8 8% | 3 3% | 10 10% |
| **HPV+, T3/T4** | 81 | 13 16% | 69 85% | 38 47% | 12 15% | 9 11% | 8 10% |
| **HPV-, T1/T2** | 43 | 2 5% | 27 63% | 11 26% | 2 5% | 1 2% | 2 5% |
| **HPV-, T3/T4** | 53 | 6 11% | 42 79% | 26 49% | 9 17% | 3 6% | 10 19% |
| **contra all** | 287 | 3 1% | 51 18% | 21 7% | 7 2% | 2 1% | 6 2% |
| **T1/T2** | 150 | 0 0% | 13 9% | 4 3% | 2 1% | 1 1% | 1 1% |
| **T3/T4** | 137 | 3 2% | 38 28% | 17 12% | 5 4% | 1 1% | 5 4% |
| **HPV+** | 181 | 3 2% | 26 14% | 13 7% | 6 3% | 2 1% | 3 2% |
| **HPV-** | 96 | 0 0% | 25 26% | 8 8% | 1 1% | 0 0% | 3 3% |

**Figure 2:**
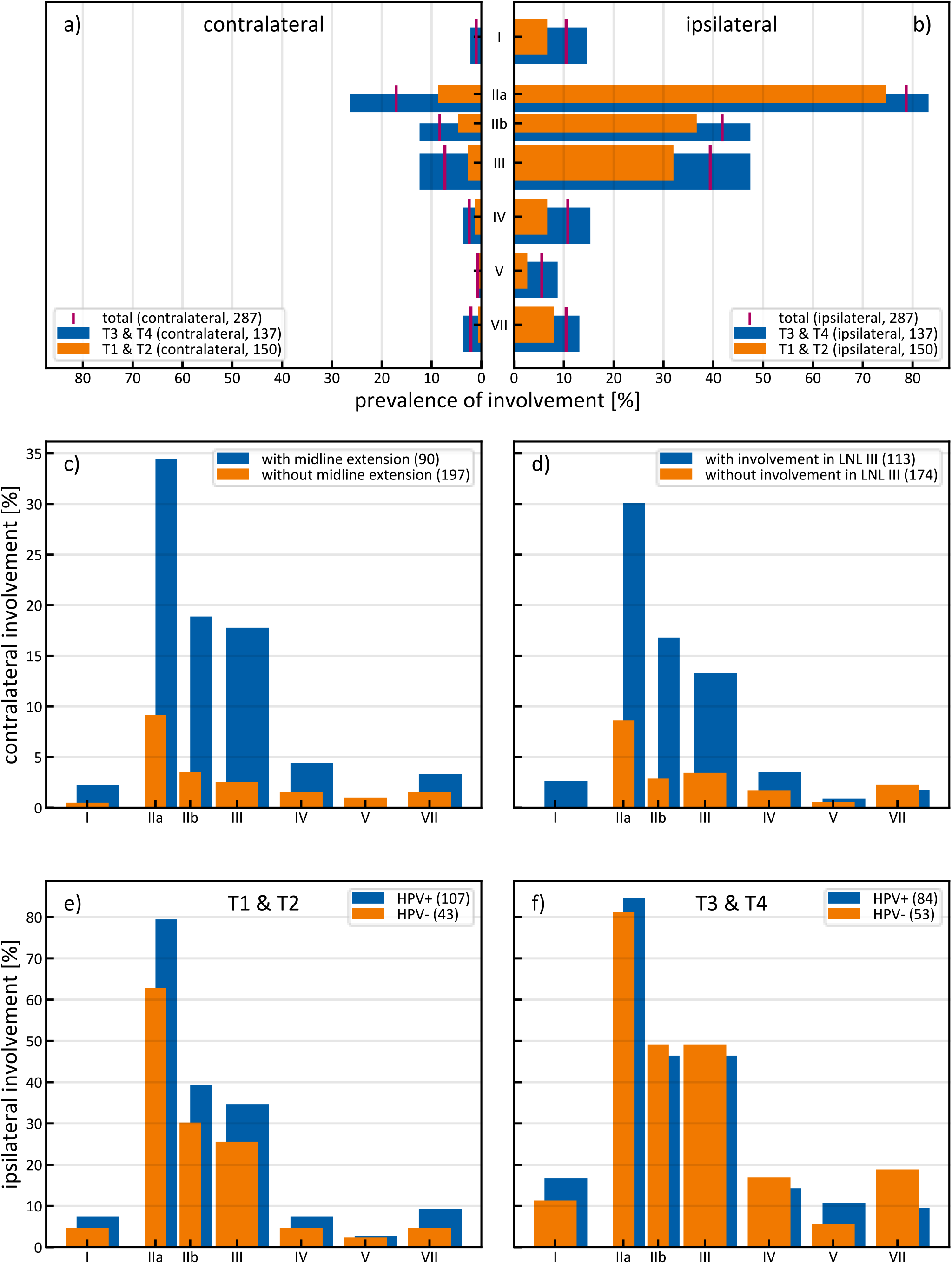
(a) Contralateral and (b) ipsilateral prevalence of LNL involvement for the whole patient cohort and stratified according to early (T1/T2) versus late (T3/T4) T-category. Contralateral LNL involvement stratified according to (c) midsagittal plan extension and (d) involvement of ipsilateral level III. Ipsilateral LNL involvement stratified according to HPV status for T1/T2 tumors (e) and T3/T4 tumors (f).

### Dependence on T-category

Table 1 and Figure 1a compares the prevalence of LNL involvement for early (T1/T2) versus late (T3/T4) T-category-patients. Consistent with common intuition, higher involvement was observed for late T-categories. Involvement of ipsilateral level II was high also for T1/T2 (79%) and therefore increased only moderately for T3/T4 (83%). However, involvement of the downstream levels III, IV, V increased from 32%, 7%, 3% for early T-category patients to 47%, 15%, 9% for late T-category. On the contralateral side, involvement of levels II, III, IV, V increased from 9%, 3%, 1%, 1% for T1/T2 patients to 28%, 12%, 4%, 1% for T3/T4.

### Dependence on upstream levels

Table 2 considers the frequency of involvement in downstream levels depending on the involvement in upstream levels. On the ipsilateral side, level III harbored metastases in 47% of patients (108 out of 232) when level II was positive, but in only 9% of patients (5 out of 55) when II was negative. Analogously, level IV harbored metastases in 25% of patients (28 out of 118) when level III was positive, but in only 2% of patients (3 out of 174) when III was negative (Figure 3). On the contralateral side, level III harbored metastases in 35% of patients (18 out of 51) when level II was positive, but in only 1% of patients (3 out of 236) when II was negative.

**Table 2:** Simultaneous involvement in levels II, III, and IV and frequency of skip metastases, for the whole patient cohort (all) and stratified according to early (T1/T2) versus late (T3/T4) T-category. Columns 1-3 define the 8 possible combinations of involvement; subsequent columns report the number of patients with the respective combination of co-involved levels.

|  |  |  | ipsilateral |  |  |  |  |  | contralateral |  |  |  |  |  |
| --- | --- | --- | --- | --- | --- | --- | --- | --- | --- | --- | --- | --- | --- | --- |
| II | III | IV | all |  | T1/T2 |  | T3/T4 |  | all |  | T1/T2 |  | T3/T4 |  |
| pos | pos | pos | 28 | 10% | 8 | 5% | 20 | 15% | 5 | 2% | 1 | 1% | 4 | 3% |
| pos | pos | neg | 80 | 28% | 37 | 25% | 43 | 31% | 13 | 5% | 2 | 1% | 11 | 8% |
| pos | neg | pos | 2 | 1% | 2 | 1% | 0 | 0% | 0 | 0% | 0 | 0% | 0 | 0% |
| pos | neg | neg | 122 | 43% | 71 | 47% | 51 | 37% | 33 | 11% | 10 | 7% | 23 | 17% |
| neg | pos | pos | 0 | 0% | 0 | 0% | 0 | 0% | 1 | 0% | 1 | 1% | 0 | 0% |
| neg | pos | neg | 5 | 2% | 3 | 2% | 2 | 1% | 2 | 1% | 0 | 0% | 2 | 1% |
| neg | neg | pos | 1 | 0% | 0 | 0% | 1 | 1% | 1 | 0% | 0 | 0% | 1 | 1% |
| neg | neg | neg | 49 | 17% | 29 | 19% | 20 | 15% | 232 | 81% | 136 | 91% | 96 | 70% |
|  |  |  | 287 |  | 150 |  | 137 |  | 287 |  | 150 |  | 137 |  |

**Figure 3:**
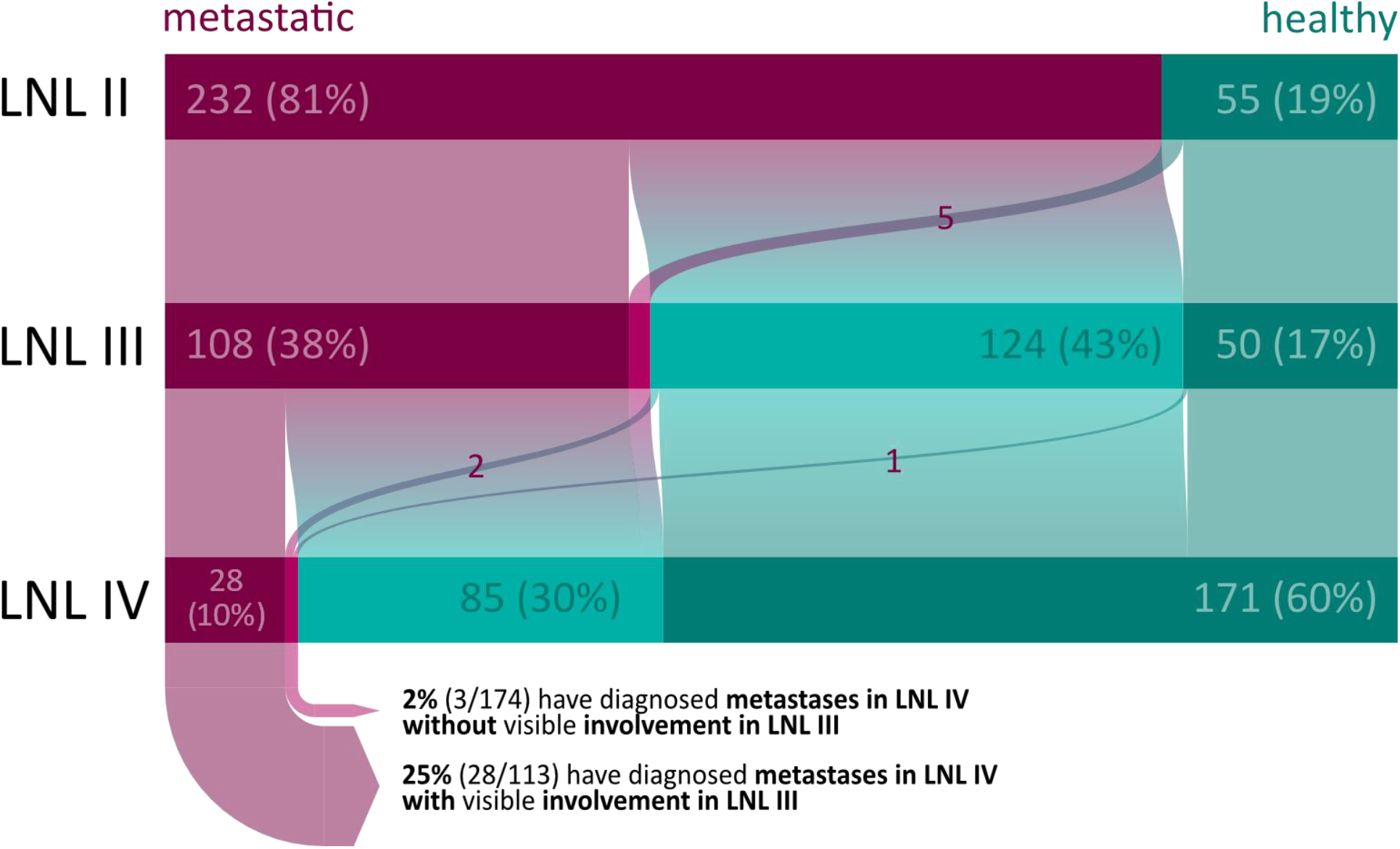
Ipsilateral involvement in levels III and IV depending on the involvement of upstream levels as flow plot.

### Contralateral involvement

Apart from late T-category (Table 1), extension of the primary tumor across the midsagittal plane and higher ipsilateral involvement was associated with higher contralateral involvement. Table 3 reports the prevalence of contralateral lymph node involvement depending on three factors: T-category, midsagittal extension, and whether ipsilateral level III was involved. For all 197 patients without midline extension, contralateral involvement in levels II, III, IV, V was 10%, 3%, 2%, 1% compared to 36%, 18%, 4%, 0% with midline extension (90 patients). In addition, out of 38 patients with late T-category, midsagittal extension, and positive ipsilateral level III, 21 (55%) showed contralateral involvement in level II and 12 (32%) in level III. Out of 39 late T-category patients with midsagittal extension but negative ipsilateral level III, contralateral involvement was lower (24% in level II, 7% in level III). In Table 3, we consider ipsilateral level III rather than II, because level II is involved in 81% of all patients. However, when ipsilateral level II is not involved, contralateral involvement is unlikely (1 out of 55 patients showed metastases in contralateral level II). We further note that the three factors considered in Table 3 are correlated. Out of 150 early T-category patients, 11 (7%) showed midline extension, whereas 79 (58%) out of 137 late T-category patients showed midline extension. As expected, contralateral involvement depended on primary tumor subsite. When considering only primary tumors strictly restricted to the tonsils (118 patients), contralateral involvement in levels II and III was 8% and 3%, respectively.

**Table 3:** Risk factors for contralateral involvement. Columns 1-3 define the 8 possible combinations of positive/negative mid-sagittal plan extension, late/early T-category, and positive/negative involvement of ipsilateral level III. Subsequent columns report the number of patients and percentages with involvement in the respective level for each combination of risk factors.

| T-cat. | midline | ipsi III | n | I |  | II |  | III |  | IV |  | V |  | VII |  |
| --- | --- | --- | --- | --- | --- | --- | --- | --- | --- | --- | --- | --- | --- | --- | --- |
| early | no | neg | 94 | 0 | 0% | 4 | 4% | 2 | 2% | 1 | 1% | 0 | 0% | 1 | 1% |
| early | no | pos | 45 | 0 | 0% | 8 | 18% | 1 | 2% | 1 | 2% | 1 | 2% | 0 | 0% |
| early | yes | neg | 8 | 0 | 0% | 0 | 0% | 0 | 0% | 0 | 0% | 0 | 0% | 0 | 0% |
| early | yes | pos | 3 | 0 | 0% | 1 | 33% | 1 | 33% | 0 | 0% | 0 | 0% | 0 | 0% |
| late | no | neg | 31 | 0 | 0% | 3 | 10% | 1 | 3% | 1 | 3% | 1 | 3% | 2 | 6% |
| late | no | pos | 27 | 1 | 4% | 4 | 15% | 1 | 4% | 0 | 0% | 0 | 0% | 0 | 0% |
| late | yes | neg | 41 | 0 | 0% | 10 | 24% | 3 | 7% | 1 | 2% | 0 | 0% | 1 | 2% |
| late | yes | pos | 38 | 2 | 5% | 21 | 55% | 12 | 32% | 3 | 8% | 0 | 0% | 2 | 5% |
|  |  |  | 287 | 3 |  | 51 |  | 21 |  | 7 |  | 2 |  | 6 |  |

### Dependence on HPV status

The HPV status was positive for 181 (63%), negative for 96 (33%), and unknown for 10 patients (4%). When considering LNL involvement for all T-categories and primary tumor subsites combined, the dataset provides no strong indication that LNL involvement is different for HPV positive versus HPV negative patients, neither regarding of the patterns of spread to the LNLs nor in terms of prevalence of involvement. However, the data suggests that the association of higher lymph node involvement with more advanced T-category is more pronounced for HPV negative tumors than for HPV positive tumors, i.e., HPV-tumors tend to disseminate earlier to regional nodes (Table 1). For example, for HPV positive tumors, involvement of ipsilateral level III increased from 35% for early T-category to 47% for late T-category. Instead, for HPV negative tumors, involvement was 26% versus 49%.

### Involvement of levels I, V, and VII

Prevalence of ipsilateral involvement was 10% (30 out of 287 patients) in level I and 6% (16 out of 287 patients) in level V. No patient had metastases in ipsilateral level I or V without involvement of ipsilateral level II. Prevalence of ipsilateral level VII involvement was 10% (30 out of 287 patients) and was more frequent for tumors of the tonsil (14%, 17 out of 118 patients) than for tumors of the base-of-tongue (6%, 5 out of 83 patients). 4 patients had metastases in ipsilateral level VII without involvement of level II. Features of more advanced disease was associated with higher involvement. For example, involvement in levels I, V, VII was 6%, 1%, 6% in early T-category patient without metastases in ipsilateral level III (102 patients) and 22%, 14%, 17% for late T-category patient with metastases in ipsilateral level III (68 patients).

## Discussion

The purpose of this study was to provide detailed per-level-quantifications of cervical lymph node involvement for oropharyngeal carcinoma on a patient-individual basis, depending on T-category, involvement of other nodal levels, and various clinicopathological factors such as smoking and HPV status. To the best of our knowledge, this is the only study providing such detailed quantitative information considering multimodal diagnostic modalities, which distinguishes this study from previous publications on the overall prevalence of LNL involvement for oropharyngeal cancer. Furthermore, an elaborate user-friendly GUI is provided to visualize and explore the dataset and study the dependence of LNL involvement as a function of the above parameters. While only selected information can be presented here in the form of tables and figures, the GUI can be used to access the full information contained in the dataset and study the influence of other factors such as primary tumor subsite or smoking status.

The main limitation of this dataset is that pathological involvement for the surgically treated patients was not available because neck dissection was performed en bloc. In addition, as a single-institution dataset, the number of patients is limited. However, dataset and GUI are made publicly available. The dataset can in the future be pooled with other datasets without loss of information, and the software platform and GUI developed may serve as the basis for collecting large multi-institutional datasets.

### Comparison to previous works

Lymph node involvement observed in this study is consistent with previous publications regarding prevalence [8], [12], [13], dependence on upstream levels [16], contralateral spread [11], [17]–[20], and HPV-dependence [14]. This is discussed in more detail in Appendix B: Comparison to other works and clinical relevance.

### From macroscopic progression patterns to risk of microscopic involvement

In this work, we consider lymphatic progression patterns assessed through imaging. Hence, the distribution of macroscopic lymph node metastases is studied. For defining the elective CTV or the extent of surgical resection, we are instead interested in the conditional probability of microscopic involvement in a LNL given that no macroscopic metastases are seen in that level. This risk depends on two aspects: 1) The sensitivity and specificity of diagnostic imaging, i.e., the probability of not diagnosing lymph node metastases that are present; and 2) The probability that tumor cells have spread to a LNL, given the observed state of tumor progression for that patient. The latter can be obtained from datasets of metastatic progression patterns in a cohort of patients as presented in this paper. Statistical methods that combine both aspects to calculate risk of microscopic involvement have been developed for ipsilateral levels I-IV [21], [22]. Future work will extend these statistical models to contralateral spread and levels V and VII, informed by the data presented here. However, this is not part of the current paper.

### Summary and Prospect

Detailed datasets of lymphatic progression patterns, meaning reporting the combination of simultaneously involved LNLs together with tumor characteristics on a patient-individual basis, allows for better quantification of LNL involvement. This may in turn allow for further personalization of elective CTV-N definition based on the individual patient’s state of tumor progression. In this paper we publish such a dataset together with a graphical user interface to explore the dataset. The software tools are made publicly available for others to study our dataset in detail and to contribute data for building larger multi-institutional datasets. Large datasets, possibly containing thousands of patients, together with the statistical methods for analysis, may eventually inform future clinical trials and guidelines on nodal CTV definition in head & neck cancer. Potential applications are to omit irradiation of ipsilateral level IV in selected patients, or to identify additional patients in whom contralateral neck irradiation can be omitted or limited to level II.

## Data Availability

All data produced are available online at

https://github.com/rmnldwg/lydata/tree/main/2021-oropharynx

https://2021-oropharynx.lyprox.org/patients/download

### Appendix A: Details on data curation

We included patients with newly diagnosed oropharyngeal squamous cell carcinoma treated at the department of radiation oncology and/or head and neck surgery of the University Hospital Zurich (USZ) between 2013 and 2019. Patients with prior radiotherapy or surgery to the neck were excluded, resulting in a dataset of 287 patients. Specific subsites of oropharyngeal cancer included the base of tongue, the tonsils as well as the oropharyngeal side of the vallecula and the posterior or lateral wall of the oropharynx. Data collection was performed by 2 experienced radiation oncologists by reviewing radiology and pathology reports together with the diagnostic images.

#### Criteria for lymph node involvement

A LNL was considered involved if the main mass of at least one malignant lymph node was located within the level. Criteria for considering a lymph node as malignant followed the description in Biau et al [23] were as follows:

- CT and MRI: Lymph nodes larger than 1 cm in the smallest transverse diameter were considered positive. In addition, all lymph nodes showing central necrosis and/or loss of fatty hilum were labeled positive
- PET-CT/MR: Nodes with an SUV of 2.5 or more and fulfilling the above criteria regarding the underlying registered CT or MRI were considered positive. Additionally, nodes with an SUV of 2.5 or more not fulfilling the CT/MRI criteria above were also considered positive if not proven negative on FNA-derived cytology. Nodes without considerable FDG-uptake (SUV<2.5) were always considered negative.
- FNA: FNA was performed based on institutional practice. 235 patients had at least one node punctured. LNL with no FNA performed were labeled ‘unknown’, LNL with positive findings were labeled positive. If the FNA findings were negative, the LNL was labeled healthy, even though only selected nodes in the level were punctured. Thus, in the interpretation of FNA involvement it must be taken into account that a negative result does not exclude the possibility of occult metastases in the LNL.
- radiotherapy planning CT: A LNL was considered positive if it contained the main mass of a lymph node contoured as nodal gross tumor volume (GTV-N), and thus may be considered a consensus decision originating from the available diagnostic modalities. For patients receiving radiotherapy following neck dissection, the planning CT is labeled negative for the resected levels.

#### Pathology after neck dissection

Finally, in case patients received a neck dissection, we recorded whether the histology showed signs of positive extra-nodal extension and how many malignant lymph nodes were resected. However, neck dissection was performed en bloc and not separated by level so that detailed information on the location of pathological lymph nodes was not available. Therefore, pathology findings are not contained in the results reported in this paper.

#### Primary tumor characteristics

Tumors extending over the midline were assigned to left/right according to the main primary tumor mass. When this was not possible, tumors were defined as ‘central’, in which case the side with more lymph node involvement was defined as ipsilateral.

Specific subsites (ICD-O-3 codes) of oropharyngeal cancer included the base of tongue, the tonsils as well as the oropharyngeal side of the vallecula and the posterior or lateral wall of the oropharynx. All tumors were assigned to one ICD-O-3 code. For extended primary tumors, this was done based on the location of the main mass. For visualization in the GUI, subsites were grouped into three categories: 1) Base of tongue (C01.9, 2) Tonsil (C09.0, C09.1, C09.8, C09.9), and Other (C10.0, C10.1, C10.2, C10.3, C10.4, C10.8, C10.9). However, the detailed subsite information is contained in the csv data base file and accessible via the patient list in the GUI.

### Appendix B: Comparison to other works and clinical relevance

#### Prevalence of LNL involvement

Overall patterns and prevalence of LNL involvement in our study (Table 1) is consistent with previous studies [8], [12], [13]. Our study contained a relatively low number of N0 patients (16%) compared to previous reports, which may be explained by differences in patient selection and diagnostic modalities used. Our study includes all patients treated at our institution between 2013 and 2019 irrespective of primary treatment. Hence, our patient cohort may be considered relatively unbiased. Studies reporting on surgically treated patients may introduce bias towards lower prevalence of LNL involvement when patients with advanced disease are referred to definitive chemoradiotherapy.

#### Dependence on upstream levels

The question on how the probability of metastases in a LNL depends on the involvement of upstream levels is poorly reported in the literature. To our knowledge, Sanguinetti et al [16] is the only publication reporting on this question for early T-category surgically treated OSCC. For example, out of 42 patients with ipsilateral level III involved, 12 patients (29%) had also level IV involved^1^, which is similar to our findings (28 out of 113 patients, 25%, for all patients combined). In agreement with previous studies, our dataset confirms that skip metastases in levels III and IV occur but are rare (Table 2). Furthermore, we observed no case of level I or V involvement without metastases in level II, which is also confirming previous publications. Further data collection and analysis in that direction could potentially lead to treatment-de-escalation strategies by not irradiating down-stream LNLs in the absence of metastases in upstream levels, e.g., by identifying patients in whom level IV may be excluded from the CTV-N.

#### Contralateral spread

A prominent example of treatment de-escalation is the sparing of the elective contralateral irradiation or the pN0, negative, neck: Chronowski et al. [17] provided data of 102 patients with tonsillar carcinoma treated with unilateral radiotherapy, of which only 2% experienced contralateral recurrence. Very similar data, with contralateral recurrence rates of only 2-3.5% were reported from the Princess Margaret Hospital [18], [19] Moreover, similarly to the results of the large meta-analysis of Al-Mamgani [11], we could demonstrate that the probability of contralateral involvement also strongly depends on T-category and midline extension. Concerning omission of radiotherapy to the pN0 neck, a recent prospective trial, with most of the patients included suffering from oropharyngeal cancer, could demonstrate excellent tumor control rates of 97% in the unirradiated neck [20]. These results show that CTV-N reduction is possible for selected patients. However, according to current guidelines, unilateral radiotherapy is recommended in specific circumstances. In our study, incidence of contralateral involvement was 20% and the data suggests that for many patients with favorable characteristics (early T-category, no midline extension, limited ipsilateral involvement), the risk of contralateral metastases is low (Table 3). If supported by further multi-institutional data, this could identify additional patients in whom the contralateral neck may be safely excluded from the CTV-N, either completely or in part. E.g. radiotherapy could be limited to level II in some patients when level II still bears a relevant risk of occult metastases but the risk in levels III and IV is minimal.

#### HPV-status

Consistent with the findings of Bauwens et al [14] and the general clinical observation that HPV-positive tumors seem to metastasize early despite small primaries, our data suggests that the dependence of lymph node involvement on T-category is less pronounced in HPV-positive tumors (Table 2). Beyond that, our data does not provide evidence that lymphatic progression patterns differ between HPV-positive versus negative tumors (consistent with Bauwens et al).

## Footnotes

1 These numbers are reconstructed from the data reported but are not directly contained in the publication.

## References

[1] J. Biau et al., “Selection of lymph node target volumes for definitive head and neck radiation therapy: a 2019 Update,” Radiother Oncol, vol. 134, pp. 1–9, 2019, doi: 10.1016/j.radonc.2019.01.018.

[2] A. L. Grosu and C. Nieder, Target volume definition in radiation oncology. Springer, 2015.

[3] R. Goel, W. Moore, B. Sumer, S. Khan, D. Sher, and R. M. Subramaniam, “Clinical Practice in PET/CT for the Management of Head and Neck Squamous Cell Cancer,” AJR Am J Roentgenol, vol. 209, no. 2, pp. 289–303, Aug. 2017, doi: 10.2214/AJR.17.18301.

[4] J. Park et al., “Diagnostic Accuracy and Confidence of [18F] FDG PET/MRI in comparison with PET or MRI alone in Head and Neck Cancer,” Sci Rep, vol. 10, no. 1, p. 9490, Jun. 2020, doi: 10.1038/s41598-020-66506-8.

[5] K. Jensen et al., “Imaging for Target Delineation in Head and Neck Cancer Radiotherapy,” Semin Nucl Med, vol. 51, no. 1, pp. 59–67, Jan. 2021, doi: 10.1053/j.semnuclmed.2020.07.010.

[6] M. Rohde et al., “18F-fluoro-deoxy-glucose-positron emission tomography/computed tomography in diagnosis of head and neck squamous cell carcinoma: a systematic review and meta-analysis,” Eur J Cancer, vol. 50, no. 13, pp. 2271–2279, Sep. 2014, doi: 10.1016/j.ejca.2014.05.015.

[7] V. Gregoire et al., “Delineation of the neck node levels for head and neck tumors: a 2013 update. DAHANCA, EORTC, HKNPCSG, NCIC CTG, NCRI, RTOG, TROG consensus guidelines,” Radiother Oncol, vol. 110, no. 1, pp. 172–81, Jan. 2014, doi: 10.1016/j.radonc.2013.10.010.

[8] V. Gregoire, E. Coche, G. Cosnard, M. Hamoir, and H. Reychler, “Selection and delineation of lymph node target volumes in head and neck conformal radiotherapy. Proposal for standardizing terminology and procedure based on the surgical experience,” Radiother Oncol, vol. 56, no. 2, pp. 135–50, Aug. 2000.

[9] V. Gregoire, A. Eisbruch, M. Hamoir, and P. Levendag, “Proposal for the delineation of the nodal CTV in the node-positive and the post-operative neck,” Radiother Oncol, vol. 79, no. 1, pp. 15–20, Apr. 2006, doi: 10.1016/j.radonc.2006.03.009.

[10] S. V. Bratman et al., “CCTG HN.10: A phase II single-arm trial of elective volume adjusted de-escalation radiotherapy (EVADER) in patients with low-risk HPV-related oropharyngeal squamous cell carcinoma (NCT03822897).,” JCO, vol. 38, no. 15_suppl, pp. TPS6592–TPS6592, May 2020, doi: 10.1200/JCO.2020.38.15_suppl.TPS6592.

[11] A. Al-Mamgani et al., “Contralateral regional recurrence after elective unilateral neck irradiation in oropharyngeal carcinoma: A literature-based critical review,” Cancer Treat Rev, vol. 59, pp. 102–108, Sep. 2017, doi: 10.1016/j.ctrv.2017.07.004.

[12] F. C. Candela, K. Kothari, and J. P. Shah, “Patterns of cervical node metastases from squamous carcinoma of the oropharynx and hypopharynx,” Head Neck, vol. 12, no. 3, pp. 197–203, Jun. 1990, doi: 10.1002/hed.2880120302.

[13] Z. Iyizoba-Ebozue et al., “Retropharyngeal Lymph Node Involvement in Oropharyngeal Carcinoma: Impact upon Risk of Distant Metastases and Survival Outcomes,” Cancers (Basel), vol. 12, no. 1, p. E83, Dec. 2019, doi: 10.3390/cancers12010083.

[14] L. Bauwens et al., “Prevalence and distribution of cervical lymph node metastases in HPV-positive and HPV-negative oropharyngeal squamous cell carcinoma,” Radiother Oncol, vol. 157, pp. 122–129, Apr. 2021, doi: 10.1016/j.ra-donc.2021.01.028.

[15] Django. Lawrence, Kansas: Django Software Foundation, 021. [Online]. Available: https://www.djangoproject.com/

[16] G. Sanguineti et al., “Defining the risk of involvement for each neck nodal level in patients with early T-stage node-positive oropharyngeal carcinoma,” International journal of radiation oncology, biology, physics, vol. 74, no. 5, pp. 1356–64, Aug. 2009, doi: 10.1016/j.ijrobp.2008.10.018.

[17] G. M. Chronowski et al., “Unilateral radiotherapy for the treatment of tonsil cancer,” Int J Radiat Oncol Biol Phys, vol. 83, no. 1, pp. 204–209, May 2012, doi: 10.1016/j.ijrobp.2011.06.1975.

[18] S. H. Huang et al., “Re-evaluation of Ipsilateral Radiation for T1-T2N0-N2b Tonsil Carcinoma at the Princess Margaret Hospital in the Human Papillomavirus Era, 25 Years Later,” Int J Radiat Oncol Biol Phys, vol. 98, no. 1, pp. 159–169, May 2017, doi: 10.1016/j.ijrobp.2017.01.018.

[19] B. O’Sullivan et al., “The benefits and pitfalls of ipsilateral radiotherapy in carcinoma of the tonsillar region,” Int J Radiat Oncol Biol Phys, vol. 51, no. 2, pp. 332–343, Oct. 2001, doi: 10.1016/s0360-3016(01)01613-3.

[20] J. A. Contreras et al., “Eliminating Postoperative Radiation to the Pathologically Node-Negative Neck: Long-Term Results of a Prospective Phase II Study,” J Clin Oncol, vol. 37, no. 28, pp. 2548–2555, Oct. 2019, doi: 10.1200/JCO.19.00186.

[21] B. Pouymayou, P. Balermpas, O. Riesterer, M. Guckenberger, and J. Unkelbach, “A Bayesian network model of lymphatic tumor progression for personalized elective CTV definition in head and neck cancers,” Phys Med Biol, vol. 64, no. 16, p. 165003, 14 2019, doi: 10.1088/1361-6560/ab2a18.

[22] R. Ludwig, B. Pouymayou, P. Balermpas, and J. Unkelbach, “A hidden Markov model for lymphatic tumor progression in the head and neck,” Sci Rep, vol. 11, no. 1, p. 12261, Jun. 2021, doi: 10.1038/s41598-021-91544-1.

[23] J. Biau et al., “Selection of lymph node target volumes for definitive head and neck radiation therapy: a 2019 Update,” Radiotherapy and Oncology, vol. 134, pp. 1–9, May 2019, doi: 10.1016/j.radonc.2019.01.018.

